# Biases in Race and Ethnicity Introduced by Filtering Electronic Health Records for ‘Complete Data’

**DOI:** 10.1101/2024.10.04.24314914

**Authors:** Jose M. Acitores Cortina, Yasaman Fatapour, Michael Zietz, Kathleen LaRow Brown, Undina Gisladottir, Danner Peter, Oliver John Bear Don’t Walk, Aditi Kuchi, Apoorva Srinivasan, Hongyu Liu, Jacob Berkowitz, Kevin Tsang, Nadine Friedrich, Sophia Kievelson, Nicholas P. Tatonetti

**Author notes:** Correspondence: 8687 Melrose Ave, West Hollywood, CA 90069, USA. Equal contribution, listed alphabetically (Jose M. Acitores Cortina, Yasaman Fatapour). Equal contribution, listed alphabetically (Michael Zietz, Kathleen LaRow Brown, Undina Gisladottir). Equal contribution, listed alphabetically (Danner Peter, Oliver John Bear Don’t Walk). Equal contribution, listed alphabetically (Aditi Kuchi, Apoorva Srinivasan, Hongyu Liu, Jacob Berkowitz, Kevin Tsang, Nadine Friedrich, Sophia Kievelson).

## Abstract

**Objective:** Integrated clinical databases from national biobanks have advanced the capacity for disease research. Data quality and completeness filters are used when building clinical cohorts to address limitations of data missingness. However, these filters may unintentionally introduce systemic biases when they are correlated with race and ethnicity. In this study, we examined the race/ethnicity biases introduced by applying common filters to four clinical records databases.

**Materials and Methods:** We used 19 filters commonly used in electronic health records research on the availability of demographics, medication records, visit details, observation periods, and other data types. We evaluated the effect of applying these filters on self-reported race and ethnicity. This assessment was performed across four databases comprising approximately 12 million patients.

**Results:** Applying the observation period filter led to a substantial reduction in data availability across all races and ethnicities in all four datasets. However, among those examined, the availability of data in the white group remained consistently higher compared to other racial groups after applying each filter. Conversely, the Black/African American group was the most impacted by each filter on these three datasets, Cedars-Sinai dataset, UK-Biobank, and Columbia University Dataset.

**Discussion and Conclusion:** Our findings underscore the importance of using only necessary filters as they might disproportionally affect data availability of minoritized racial and ethnic populations. Researchers must consider these unintentional biases when performing data-driven research and explore techniques to minimize the impact of these filters, such as probabilistic methods or the use of machine learning and artificial intelligence.

## INTRODUCTION

The widespread adoption of electronic health records (EHRs) over the past decade has significantly increased the availability and accessibility of electronic clinical data. This advancement enables healthcare professionals to harness vast amounts of information, driving medical research, personalized medicine, and overall improvements in healthcare delivery(1,2). Additionally, it helps to build big data in healthcare, providing a foundation for advanced analytics and informed decision-making on a large scale. This supports the training and validation of AI methodologies and models, leading to improved diagnostic accuracy, personalized treatment plans, and more efficient healthcare delivery. Therefore, the collected data significantly influences the results and hypotheses derived from these methods.

Diversity in healthcare studies includes populations underrepresented in the biomedical, clinical, behavioral, and social sciences, such as individuals from racial and ethnic minority groups, those with disabilities, and people from disadvantaged backgrounds(3). Fostering diversity is vital for producing more accurate, inclusive research outcomes that reflect the needs of all populations, ultimately leading to more equitable healthcare and improved patient outcomes(4,5).

However, these available clinical data face significant challenges that limit their effectiveness and reliability across different populations, particularly when focusing on the diverse U.S. population. This lack of diversity in data points, which may be due to systemic biases and discrimination against individuals and groups from minoritized populations can lead to biased research outcomes that exacerbate health disparities(6–9). This can lead to inaccurate conclusions about treatment or interventions that may not apply equally across different populations. In addition to that, there are still not enough big clinical longitudinal datasets, which are essential for understanding long-term health trends, progression of diseases over time and evaluating treatment outcomes.

When conducting observational clinical data analysis, it is natural to aim for a dataset as complete as possible. However, well-meaning filters to ensure completeness may introduce unintended biases in the target population(10).

In this study, we aim to evaluate the effect of data completeness filters on different datasets and how various filters impact the patient cohort. Specifically, we examined race and ethnicity biases(11) introduced by applying common filters to four distinct databases, including All of Us, UK Biobank, and two geographically distinct academic medical centers.

## MATERIALS AND METHODS

We examined four distinct data sources, All of Us, UK Biobank, Columbia University dataset and Cedars-Sinai dataset comprising approximately 12 million patients. By analyzing the available data and applying each filter, we aimed to investigate the potential biases these filters may introduce.

### All of us

The All of Us study, sponsored by the National Institutes of Health, has enrolled more than 814,000 participants as of June 18th, 2024, with 80% of them coming from underrepresented populations(12). These groups include racial and ethnic minorities, people with disabilities, those in rural or underserved areas, and individuals from lower socioeconomic backgrounds. Figure A1 (provided by the All of Us study) showcases the self-reported races and ethnicities of the participants who have completed the initial steps of the program, providing a diverse representation. The recruitment process spans all regions of the United States.

The All of Us workbench encompasses a wealth of information gathered from electronic health records, including data from Fitbit devices, survey responses, and socioeconomic factors. Notably, a recent release of data in April 2023 included approximately 245,400 whole genome sequencing records and 312,940 genotyping microarrays, further enhancing the dataset’s depth and potential for analysis.

### UK biobank

The UK Biobank is a large-scale population-based study that aims to improve the prevention, diagnosis, and treatment of various diseases. It involves the collection of extensive health-related data, including genetic information, from over 500,000 participants in the United Kingdom. Participants in the UK Biobank, recruited at ages 40-69, were registered with the National Health Service (NHS). Researchers can download the data through the UK Biobank’s Data Showcase, which collaborates closely with the European Genome Archive (EGA).

### Cedars Sinai

Cedars Sinai Medical Center is one of the largest hospitals in California, based in Los Angeles, serves up to 1 million diverse patients every year across its 40 locations in southern California. CSMC also serves as a large research center. The studied database comprises over 4 million patients.

### Columbia

Columbia University Irving Medical Center is a clinical, research, and educational enterprise located on a campus in northern Manhattan. They are home to four colleges and schools that work on scientific research, education, and patient care. The studied database comprises over 5 million patients.

The self-reported race and ethnicity distributions of each dataset, along with the number of participants in each dataset, are presented in Table 1. In addition to those self-reported categories for race and ethnicity we defined an all group, which includes every patient in that specific dataset. This group serves as a baseline for comparison within the dataset.

**Table 1.**
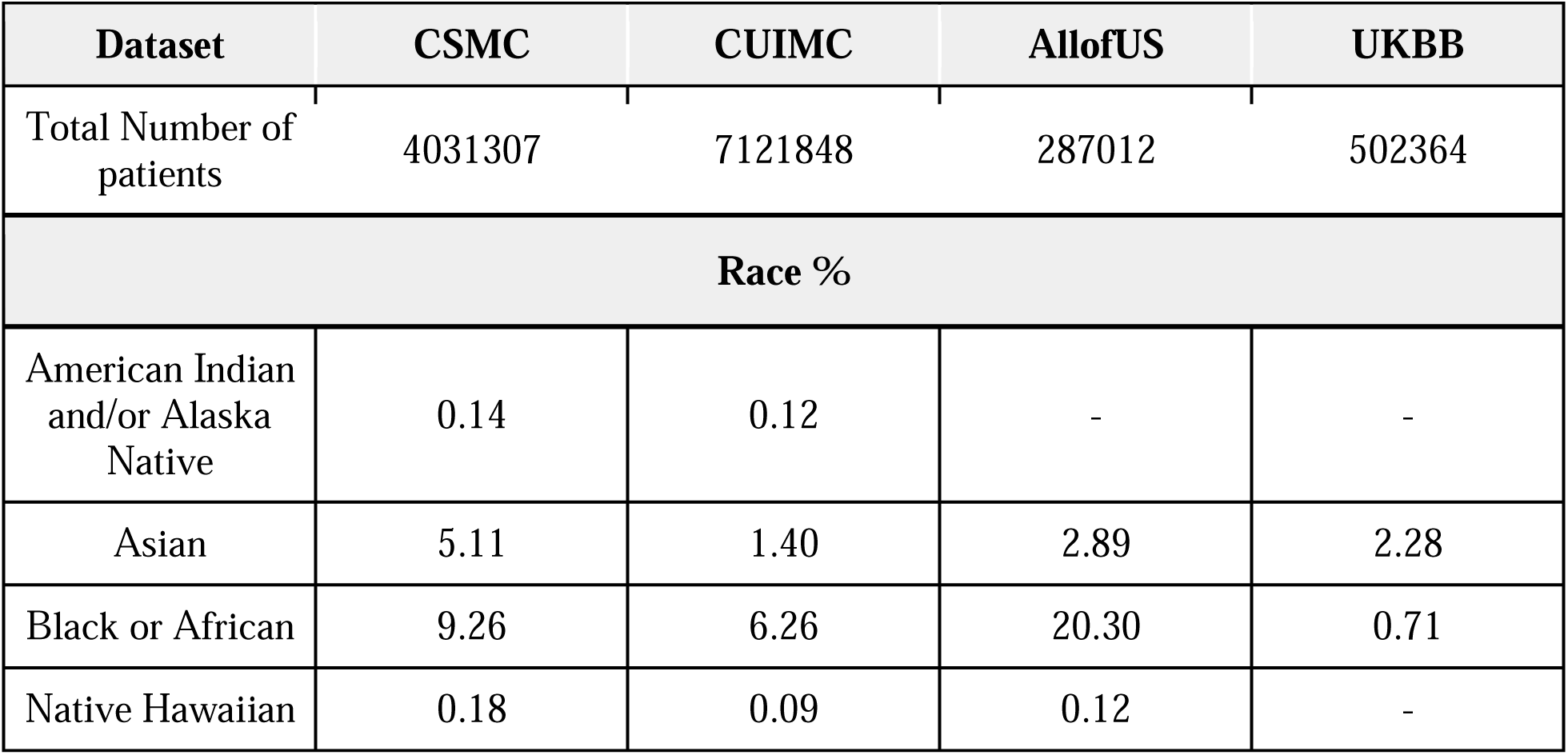
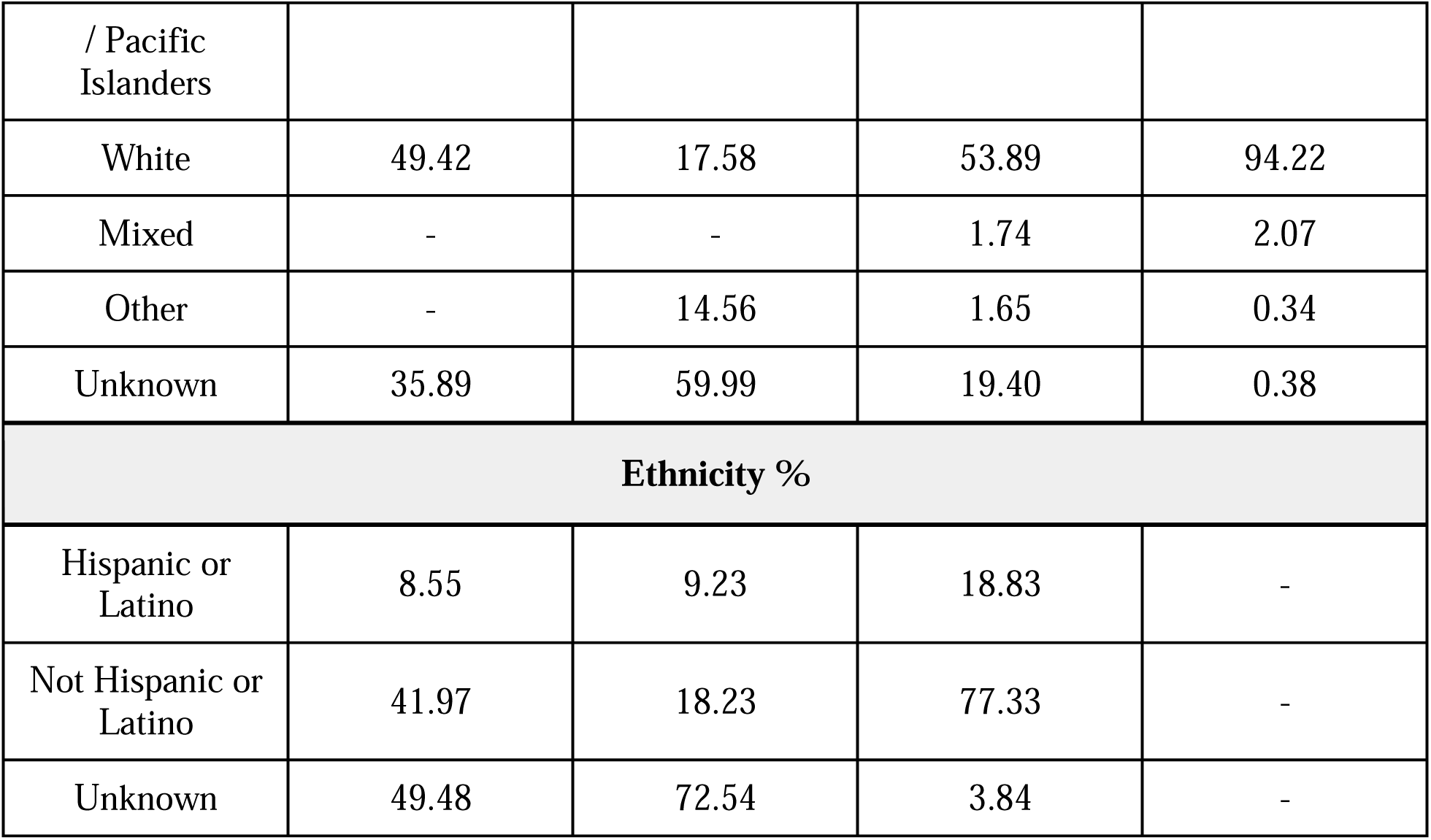
Self-reported race and ethnicity percentages of each dataset, along with the total number of participants in each dataset.

### Filters

A straightforward approach to identifying subsets of patients whose data are suitable for research studies is to use heuristic computational filters that exclude patients lacking various types of data in their records. For this study, we evaluated 19 different filters, which can be grouped into three categories. The first category is based on patient demographics. This includes filters that check whether the patient has both age and sex recorded (AgeSex), if the patient is alive at the time of the search (Alive), if the patient has a known address or zip code (Address/ZIP), and a set of age filters. The age filters have been applied to age at the time of any diagnosis, for example, the age filter >= 65 selects patients that are 65 or above 65 years old at the time of any of their recorded diagnoses.

The second category is a record-based filter, which checks whether patients have at least one recorded instance of various medical data. These filters are the presence of at least one diagnosis, the presence of medication records, and records for outpatient visits.

The last category is the time span or observational period filter, which selects patients who have had multiple interactions with the healthcare system during a specific period of time, the maximum time window for this category was the 6-year follow-up.

We used 19 filters, originally defined by Weiskopf as a metric for evaluating the completeness of EHRs, to build patient cohorts within each dataset(13). These filters helped identify the types of data available after their application. To maintain consistency across datasets, we applied these filters to the patient populations with EHR data in each dataset. Detailed descriptions of each filter along with their categories are presented in Table 2.

**Table 2.** Filters used in our analysis, grouped by category and descriptions of each filter.

| Filters and definitions |  |  |
| --- | --- | --- |
| Group | Filter | Description |
| Demographics | Alive | Patient is alive at the time of the query |
|  | AgeSex | Patient has both sex and age recorded |
| | Age filter $\geq 18$ | Patient has a diagnosis at an age included in the filter |
| | Age filter $\leq 21$ | |
| | Age filter $\leq 40$ | |
| | Age filter $\leq 65$ | |
| | Age filter $\geq 65$ | |
| | Age filter $\leq 80$ | |
|  | Address/zip code | Patient has an address or zip code recorded |
| Medical interactions | Diagnosis | Patient has at least one diagnosis recorded |
|  | Medication | Patient has at least one medication prescribed and recorded |
|  | Out-patient visit | Patient has at least one out-patient visit recorded |
| Observation period | Obs-period 1 week | Patient has a recorded observation period equal or longer than the filter span |
|  | Obs-period 2 week |  |
|  | Obs-period 1 month |  |
|  | Obs-period 6 months |  |
|  | Obs-period 1 year |  |
|  | Obs-period 2 years |  |
|  | Obs-period 6 years |  |

This study aimed to identify the biases of different types of filters which are used by researchers to evaluate data completeness in electronic EHR datasets. Our focus is biases that may be introduced upon applying these filters to races and ethnicities.

## RESULTS

We applied the filters to each dataset separately to assess their individual effects. Table 3 indicates the percentage of patients remaining after applying each filter for each dataset.

**Table 3.** Percentage of population remaining on each dataset after applying the different filters.

| Filter | Cedars-Sinai | Columbia | All of Us | UKBB |
| --- | --- | --- | --- | --- |
| Alive | 95.563 | 95.127 | 98.88 | 93.027 |
| Age/Sex | 99.870 | 84.282 | 100 | 100 |
| Age filter $\geq 18$ | 40.308 | 60.193 | 88.49 | 45.776 |
| Age filter $\leq 21$ | 3.169 | 18.652 | 14.56 | 16.513 |
| Age filter $\leq 40$ | 18.187 | 39.515 | 22.76 | 35.533 |
| Age filter $\leq 65$ | 33.438 | 61.985 | 65.16 | 44.495 |
| Age filter $\geq 65$ | 10.537 | 16.434 | 28.55 | 24.350 |
| Age filter $\leq 80$ | 39.305 | 70.099 | 84.62 | 45.776 |
| Has address/zip | 45.377 | 56.504 | 99.99 | 29.513 |
| Has medications | 32.663 | 35.296 | 83.51 | 77.373 |
| Has diagnoses | 41.263 | 72.426 | 88.66 | 92.957 |
| Has out. visits | 73.254 | 40.635 | 99.72 | 45.799 |
| Obs period 1 week | 55.636 | 70.114 | 1.83 | 44.469 |
| Obs period 2 weeks | 54.336 | 69.333 | 1.71 | 44.430 |
| Obs period 1 month | 52.636 | 68.183 | 1.53 | 44.373 |
| Obs period 6 months | 46.864 | 59.543 | 1.02 | 44.192 |
| Obs period 1 year | 43.036 | 57.328 | 1.0 | 44.056 |
| Obs period 2 years | 38.238 | 54.482 | 1.0 | 43.853 |
| Obs period 6 years | 18.188 | 44.251 | 1.0 | 43.247 |

Figure 1 shows the percentage of available patients in the Cedars-Sinai Medical Center (CSMC) after applying each filter. The results show that both unknown race and unknown ethnicity are the most affected groups when applying the filters. This causes the values for the group all to decrease too, and it’s shown across every cohort.

The results of the Cedars-Sinai Medical Center cohort show every known race/ethnicity group is above ‘all’ in almost every filter. However, both unknown race and unknown ethnicity are the most affected groups when applying the filters. This causes the values for the group all to decrease too, and it’s shown across every cohort.

Nevertheless, it is important to note that in this dataset the Black or African American population is the most affected group by the filters, being the most affected in CSMC and UKBB, the second most affected in CUIMC and the most affected or second most affected by every filter in AllofUs, this can be seen in the appendix tables A2, A3, A4, and A5. The most complete group varies depending on the filter. In the ethnicity groups we see Hispanic or Latino as the most affected one by the filters.

### All of Us

For the All of Us dataset, we applied the filters to the cohort of patients in the controlled tier 7 who had EHR records. This process reduced the number of patients from 410,235 to 287,012. Upon applying age/sex, medication, zip code, or address (in this dataset we have state of residence, so we used that instead of zip code), alive status, and outpatient visits, the initial cohort remained largely unchanged. However, as more stringent age filters were applied and the observational period was extended, the cohort population significantly decreased. Among all the races, the Asian group was most noticeably impacted, particularly when the observation period filter was applied, as shown in Figure 2. Within this dataset, unlike at CSMC, the majority of racial and ethnic groups follow the same pattern when each filter is applied.

### Columbia University Irvine Medical Center

Similarly to the cohort from CSMC, the unknown race/ethnicity and other values decrease the most when applying the filters, bringing down the overall percentage. The known races/ethnicities are again above the all-group’s percentage in almost every category. It is important to note that unknown race and ethnicity represent more than 60% and 70% of Columbia University Medical Center’s cohort respectively, contributing to the low baseline percentage for the all group.

Out of the known races and ethnicities, we can see in Figure 3 that the American Indian and/or Alaska Native population is the most affected by the filters, 17 out of 19, even crossing the all line. Followed by Black or African American which takes the second place in 10 out of 19. However, contrary to the CSMC cohort, the Not Hispanic or Latino ethnicity is the most affected by the filters, 17 out 19 filters.

### UK Biobank

In UKBB the race and ethnicity groups are different to the American institutions, we implemented the grouping strategy that the UK government provides as a guideline(14), this conversion keeps the graphs and data consistent as it follows a similar fashion as the US system; We included any other Black background, African, Black or Black British, and Caribbean, under the category Black or African origin; any other Asian background, Asian or Asian British, Bangladeshi, Chinese, Indian and, Pakistani under the category Asian; any other white background, British, Irish, white under the category white; do not know and, prefer not to answer under unknown, any other mixed background, mixed, white and Asian, white and Black African and, white and Black Caribbean under mixed and, finally, other ethnic group under other.

After this grouping, there are some aspects to remark on from this dataset, white population represents close to 94% of the group, which biases completely the all results. Having that in mind, we can see in Figure 4 how every other race was impacted more than the baseline, especially the Black or African origin group.

We then analyzed the differences within the most prevalent groups in this dataset, evaluating only the five most common categories. We found that the British group accounts for 91% only counting the top five groups. This approach yielded similar results as the complete one. The first five categories, in order of percentage were the following: British 91.7%, any other white background 3.4%, Irish 2.8%, Indian 1.2% and other ethnic groups 1%. These percentages account for the addition of the population of the top five groups and not the total.

## DISCUSSION

Our study investigates the potential racial and ethnic biases introduced by applying common data quality and completeness filters in clinical research databases, including All of Us, UK Biobank, and two academic medical centers. We analyzed 19 filters across approximately 12 million patients and discovered that certain filters significantly reduce data availability and have a differential effect on racial and ethnic groups.

The challenge with these filters lies in distinguishing between patients with missing data and those who are relatively healthy, have not recently sought medical care, or have limited access to healthcare systems. All these three groups will have a low number of data entries in their records. Consequently, these filters might bias the resulting cohort by selecting sicker patients who interact with the healthcare system more frequently and/or those who have more access to healthcare systems. For example, in a cohort of 10,000 patients, those with poorer health status had more lab tests and medication orders, resulting in more comprehensive data in their records(15). On the other hand, minoritized populations usually have less access to healthcare(16), which affects the data’s completeness and reduces their data points when we apply different types of filters. We focused on bringing attention to the second point.

Throughout the analysis of the four different cohorts, a consistent pattern emerged: applied filters disproportionately affected minoritized groups, particularly the Black/African American group, which consistently has one of the lowest data availabilities across all datasets, and the American Indian and/or Alaska Native group. These filters significantly reduced the already limited data points for minoritized groups, further diminishing the completeness and usability of their data compared to white or non-Latino patients. We observe a similar pattern in the Hispanic or Latino group, where data availability is consistently lower in every cohort compared to the non-Hispanic or non-Latino group.

In the self-reported race groups, we observe that almost every group has less data availability than the white group, which is the largest within the known self-reported races across all datasets, except at CSMC. At CSMC, the most complete group varies by filter, alternating between Asian, Native Hawaiian or Other Pacific Islander, American Indian or Alaska Native, and white. In contrast, in the CUIMC dataset, the American Indian or Alaska Native group has the lowest data availability.

Among the four distinct datasets, only the All of Us dataset closely reflects the diversity of the US population’s, with approximately 50% of the data representing populations other than White. Upon applying different filters on this dataset, as shown in Figure 4, most groups follow the same original pattern prior to applying the filters and do not significantly deviate from the baseline, demonstrating that it is possible to achieve a diverse and complete dataset.

Dataset diversity is essential for enhancing the generalizability and inclusivity of clinical research, addressing disparities, and improving healthcare outcomes for underrepresented populations. The All of Us dataset, designed as a nationwide research program, aims to collect health data from a diverse population. In contrast, the CUIMC and CSMC datasets reflect the specific patient populations of their respective regions, leading to localized diversity compared to the national scope of the All of Us dataset. However, both are based in highly racially and ethnically diverse U.S. cities, giving them a unique advantage over other institutional-level datasets. The information for the different populations and distributions of the locations for the four datasets can be found in Table A1.

Both in the United States and the United Kingdom, the white population constitutes the majority. Minorities, as defined by the U.S. Office of Management and Budget (OMB), include racial and ethnic groups such as American Indian, Alaska Native, Asian, Black or African American, and Native Hawaiian or Pacific Islander. These groups often face health disparities, which can result in reduced access to healthcare and underrepresentation in research cohorts(17). This underrepresentation may lead to inaccurate clinical care decisions, skewed genetic associations, and suboptimal treatment strategies.

Our findings highlight the importance of carefully selecting filters to ensure equitable research outcomes, particularly for minority populations. While we do not claim these are the most frequently used filters by researchers, nor the optimal ones for selecting patients with complete data, it is essential to investigate any potential biases that may be introduced upon applying each filter before conducting research on these populations.

In addition, addressing disparities in representation is critical to creating research cohorts that accurately reflect the target population. This work underscores the challenges of achieving data completeness and proper representation for racial and ethnic populations and other minoritized groups in clinical research. Strategies to mitigate these disparities, along with careful consideration of filters, are crucial for ensuring equitable research outcomes and enhancing the inclusivity of health datasets.

Advanced methods such as AI-driven synthetic data generation, bias mitigation algorithms, and advanced statistical techniques like inverse probability weighting can help address these challenges and enhance dataset diversity. By addressing these issues, researchers can enhance the reliability and applicability of their findings, ultimately contributing to a more inclusive and fair healthcare system.

## CONCLUSION

Our findings underscore the importance of using only necessary filters as they might have consequences on the diversity and completeness of population data which particularly affects underrepresented populations. Researchers must be aware of the target population when conducting research and address these unintentional biases when performing data-driven research and the variances introduced into downstream analysis.

It is also necessary to explore techniques to minimize the impact of these filters, such as probabilistic methods or the use of machine learning and artificial intelligence to perform the most fair and just analysis possible. This may include techniques to treat the data, methods to process it or different evaluation strategies

We strive to achieve a state where the datasets accurately represent the target population of the studies and where research studies are performed on the same population that institutions serve.

## Data Availability

The data is not publicly available. All of Us data can be requested.

---

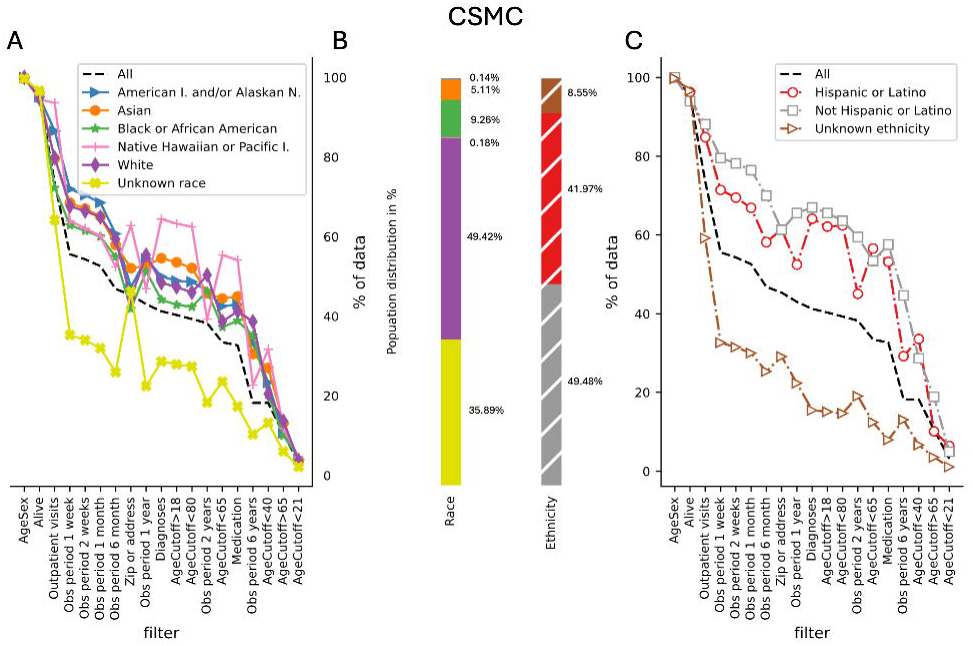

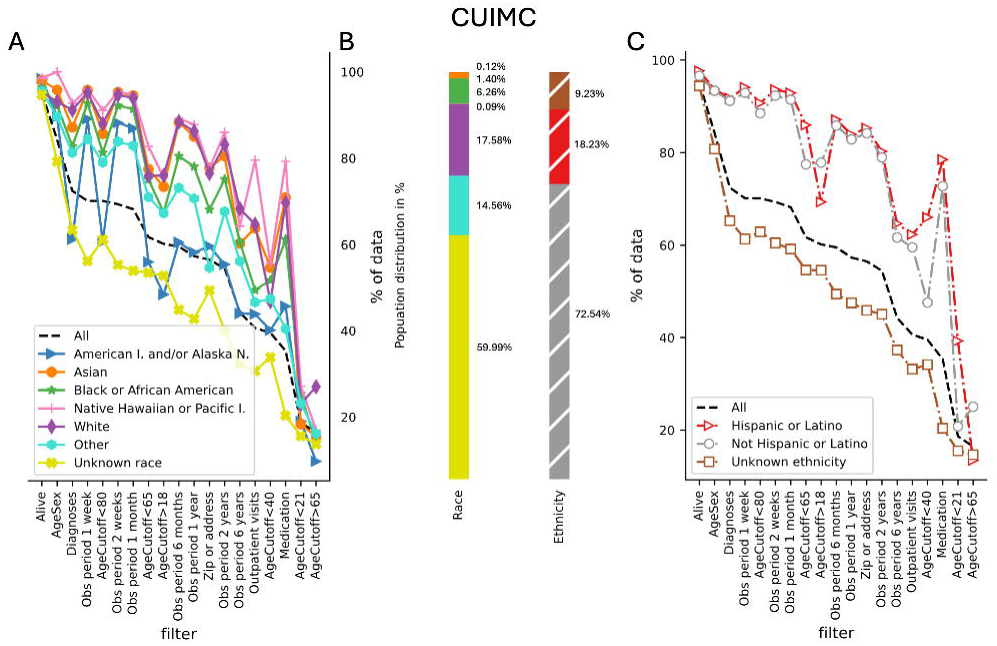

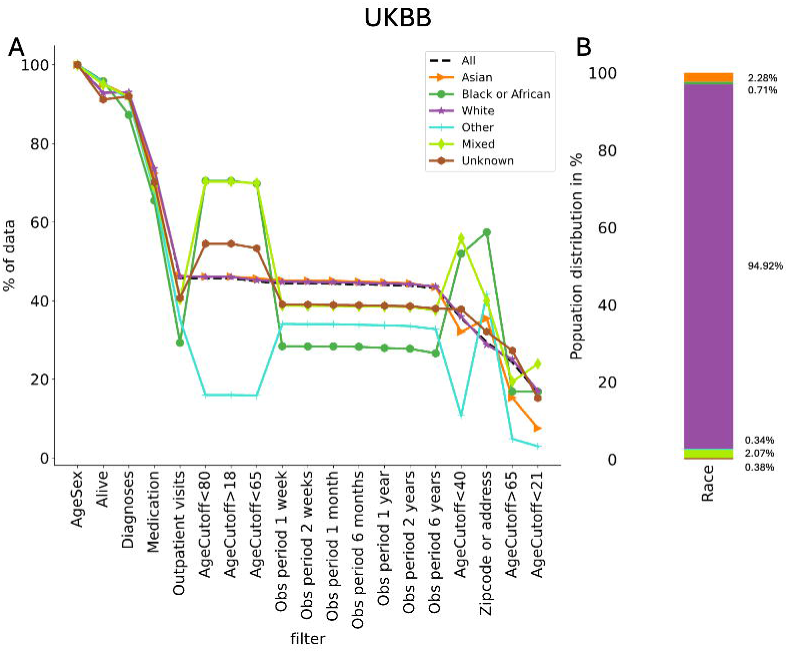

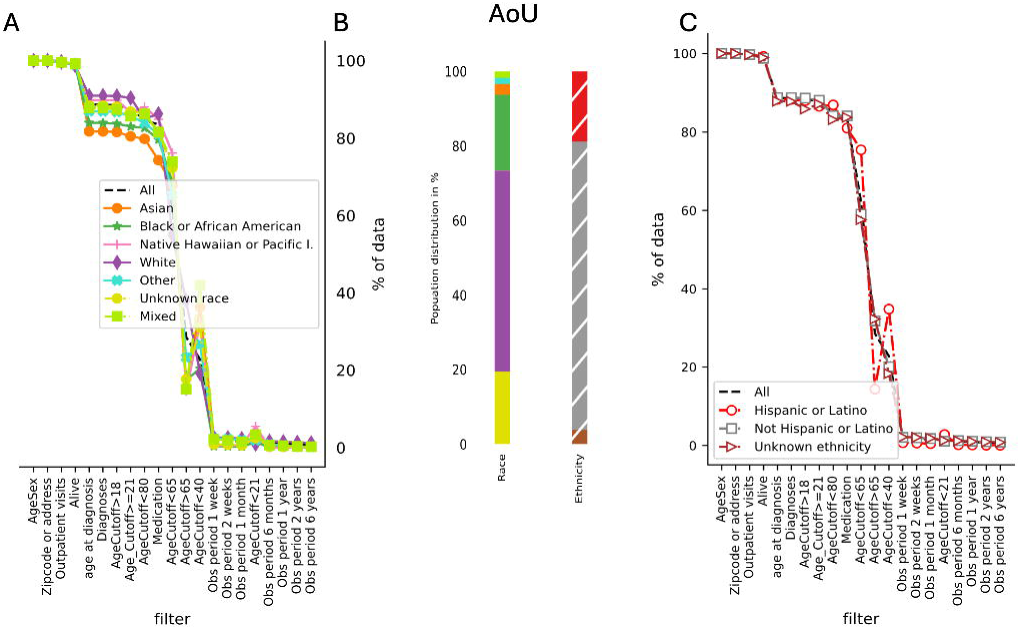

