## Supplementary material for "Biases in Race and Ethnicity Introduced by Filtering Electronic Health Records for ‘Complete Data’": Figure A

**APPENDIX**

**Table A1**

| **Dataset** | **LA County(1)** | **NY City(1)** | **US(1)** | **UK(2)** |
| --- | --- | --- | --- | --- |
| Total | 9,663,345 | 8,258,035 | 334,914,895 | 59,597,540 |
| **Race %** | | | | |
| American Indian and/or Alaska Native | 1.5 | 0.6 | 1.3 | - |
| Asian | 16.0 | 14.5 | 6.4 | 9.3 |
| Black or African | 9.0 | 23.1 | 13.7 | 4.0 |
| Native Hawaiian / Pacific Islanders | 0.4 | 0.1 | 0.3 | - |
| White | 69.6 | 37.5 | 75.3 | 81.7 |
| Mixed | 3.4 | 8.9 | 3.1 | 2.9 |
| Other | - | - | - | 2.1 |
| Unknown | - | - | - | - |
| **Ethnicity %** | | | | |
| Hispanic or Latino | 48.6 | 29.0 | 19.5 | - |
| Not Hispanic or Latino | 51.4 | 71.0 | 80.5 | - |
| Unknown | - | - | - | - |

Table A1. Self-reported race and ethnicity percentages of each location of the datasets, along with the total population number.

**Table A2**

| Filter | All | American I. and/or Alaskan N. | Asian | Black or African American | Native Hawaiian or Pacific I. | White | Unknown race | Hispanic or Latino | Not Hispanic or Latino | Unknown ethnicity |
| --- | --- | --- | --- | --- | --- | --- | --- | --- | --- | --- |
| Alive | 95.56 | 95.14 | 96.01 | 94.60 | **94.41** | 94.78 | 96.67 | 96.31 | **93.98** | 96.66 |
| AgeSex | 99.87 | 100.00 | 100.00 | 100.00 | 100.00 | 100.00 | 99.64 | 100.00 | 100.00 | 99.74 |
| AgeCutoff>18 | 40.31 | 48.90 | 53.60 | **42.86** | 63.27 | 47.36 | 27.90 | **62.11** | 65.58 | 15.11 |
| AgeCutoff<21 | 3.17 | 04.07 | **3.54** | 4.33 | 4.56 | 3.65 | 2.14 | 6.44 | **4.94** | 1.11 |
| AgeCutoff<40 | 18.19 | 23.30 | 26.96 | **20.19** | 31.64 | 20.47 | 13.20 | 33.57 | **28.65** | 6.66 |
| AgeCutoff<65 | 33.44 | 42.53 | 44.49 | **37.29** | 55.41 | 38.68 | 23.51 | 56.62 | **53.49** | 12.43 |
| AgeCutoff>65 | 10.54 | 10.64 | 13.18 | **9.98** | 11.82 | 13.64 | 06.02 | **10.15** | 18.88 | 3.53 |
| AgeCutoff<80 | 39.31 | 48.69 | 52.14 | **42.36** | 62.48 | 45.99 | 27.33 | **62.42** | 63.60 | 14.71 |
| Zip or address | 45.38 | 47.32 | 52.09 | **41.87** | 62.79 | 44.62 | 46.27 | 61.35 | **61.33** | 29.09 |
| Medication | 32.66 | 43.00 | 45.02 | **38.84** | 54.17 | 41.26 | 17.32 | **53.22** | 57.60 | 7.96 |
| Diagnoses | 41.26 | 50.04 | 54.62 | **44.17** | 64.40 | 48.43 | 28.60 | **64.12** | 66.99 | 15.49 |
| Outpatient visits | 73.52 | 86.55 | 79.62 | **72.28** | 93.65 | 79.80 | 64.18 | **84.83** | 88.11 | 59.20 |
| Obs period 1 w | 55.64 | 71.98 | 68.52 | **62.86** | 63.98 | 67.66 | 35.27 | **71.45** | 79.54 | 32.63 |
| Obs period 2 w | 54.34 | 70.43 | 67.03 | **61.47** | 62.07 | 66.40 | 33.98 | **69.49** | 78.19 | 31.49 |
| Obs period 1 m | 52.64 | 68.53 | 65.02 | **60.00** | 60.15 | 64.92 | 31.97 | **66.89** | 76.45 | 29.98 |
| Obs period 6 m | 46.86 | 60.68 | 57.98 | 54.96 | **52.46** | 59.36 | 25.91 | **58.19** | 70.00 | 25.29 |
| Obs period 1 y | 43.04 | 54.52 | 52.69 | 51.29 | **46.92** | 55.43 | 22.40 | **52.49** | 65.56 | 22.30 |
| Obs period 2 y | 38.24 | 46.39 | 45.73 | 46.32 | **39.27** | 50.39 | 18.32 | **45.07** | 59.50 | 19.03 |
| Obs period 6 y | 18.19 | 33.22 | 30.51 | 35.16 | **22.66** | 38.63 | 10.34 | **29.21** | 44.64 | 13.08 |

Table A2. Available percentage of patients' data upon applying all the 19 filters to the Cedars-Sinai Medical Center dataset. The underlined and bold text indicates the most affected known race by that filter, and the bold text indicates the most affected known ethnicity.

**Table A3**

| Filter | All | American I. and/or Alaska N. | Asian | Black or African American | Native Hawaiian or Pacific I. | White | Other race | Unknown ethnicity | Hispanic or Latino | Not Hispanic or Latino | Unknown race |
| --- | --- | --- | --- | --- | --- | --- | --- | --- | --- | --- | --- |
| Alive | 95.13 | 98.42 | 98.06 | 96.43 | 98.55 | **95.26** | 95.61 | 94.44 | 97.66 | **96.58** | 94.75 |
| AgeSex | 84.28 | **89.56** | 95.83 | 92.26 | 100.00 | 93.08 | 89.58 | 80.79 | 93.74 | **93.40** | 79.28 |
| AgeCutoff>18 | 60.19 | **48.41** | 73.42 | 67.41 | 75.54 | 76.01 | 67.24 | 54.60 | **69.35** | 77.84 | 52.78 |
| AgeCutoff<21 | 18.65 | 19.01 | **18.3**2 | 25.29 | 27.10 | 23.00 | 23.17 | 15.50 | 39.24 | **20.79** | 15.58 |
| AgeCutoff<40 | 39.52 | **40.12** | 54.66 | 51.75 | 55.58 | 46.81 | 47.39 | 34.12 | 65.98 | **47.58** | 33.81 |
| AgeCutoff<65 | 61.69 | **55.98** | 77.46 | 75.12 | 82.74 | 75.91 | 70.98 | 54.64 | 85.95 | **77.45** | 53.47 |
| AgeCutoff>65 | 16.43 | **9.69** | 15.44 | 13.92 | 16.96 | 26.98 | 16.13 | 14.66 | **13.43** | 25.02 | 13.71 |
| AgeCutoff<80 | 70.10 | **60.65** | 85.66 | 81.29 | 91.10 | 88.04 | 79.00 | 62.85 | 90.75 | **88.51** | 61.14 |
| Zip or address | 56.50 | **59.63** | 77.81 | 68.06 | 77.63 | 76.48 | 54.64 | 45.89 | 85.29 | **84.17** | 49.36 |
| Medication | 35.30 | **45.69** | 70.91 | 61.18 | 79.26 | 69.66 | 40.43 | 20.39 | 78.49 | **72.75** | 20.35 |
| Diagnoses | 72.43 | **61.23** | 87.16 | 82.78 | 92.75 | 91.20 | 81.27 | 65.25 | 91.75 | **91.19** | 63.34 |
| Outpatient visits | 40.64 | **43.87** | 63.74 | 49.42 | 79.59 | 64.66 | 46.59 | 33.13 | 62.28 | **59.55** | 30.62 |
| Obs period 1 w | 70.11 | **89.12** | 95.81 | 92.75 | 96.01 | 95.22 | 84.44 | 61.32 | 94.12 | **92.95** | 56.24 |
| Obs period 2 w | 69.33 | **88.25** | 95.36 | 92.18 | 95.49 | 94.69 | 83.85 | 60.46 | 93.63 | **92.33** | 55.31 |
| Obs period 1 m | 68.18 | **86.92** | 94.53 | 91.43 | 94.71 | 93.94 | 82.93 | 59.19 | 92.90 | **91.45** | 53.93 |
| Obs period 6 m | 59.54 | **60.52** | 88.29 | 80.49 | 89.23 | 88.44 | 73.15 | 49.42 | 87.12 | **85.85** | 44.86 |
| Obs period 1 y | 57.33 | **58.19** | 85.04 | 78.05 | 87.79 | 86.23 | 70.65 | 47.53 | 83.90 | **82.86** | 42.76 |
| Obs period 2 y | 54.48 | **55.42** | 80.49 | 75.13 | 86.00 | 83.15 | 67.64 | 45.05 | 80.24 | **78.96** | 40.07 |
| Obs period 6 y | 44.25 | **43.99** | 60.27 | 60.61 | 64.25 | 68.27 | 56.15 | 37.28 | 64.64 | **61.66** | 32.21 |

Table A3. Available percentage of patients' data upon applying all the 19 filters to the Columbia University Irving Medical Center dataset. The underlined and bold text indicates the most affected known race by that filter, and the bold text indicates the most affected known ethnicity.

**Table A4**

| Filter | All | Asian | Black or African American | Mixed | Native Hawaiian or Pacific I. | White | Other | Unknown race | Hispanic or Latino | Not Hispanic or Latino | Unknown ethnicity |
| --- | --- | --- | --- | --- | --- | --- | --- | --- | --- | --- | --- |
| Alive | 98.88 | 99.28 | 98.75 | 99.20 | **98.55** | 98.78 | 98.98 | 99.21 | **99.25** | 98.79 | 98.95 |
| AgeSex | 100.00 | 100.00 | 100.00 | 100.00 | 100.00 | 100.00 | 100.00 | 100.00 | 100.00 | 100.00 | 100.00 |
| AgeCutoff>18 | 88.49 | **81.59** | 83.70 | 87.40 | 89.83 | 90.89 | 86.74 | 88.13 | **88.00** | 88.65 | 85.87 |
| AgeCutoff<21 | 1.42 | 2.19 | 1.41 | 3.34 | 5.52 | **0.93** | 1.88 | 2.46 | 2.80 | **1.09** | 1.26 |
| AgeCutoff<40 | 22.76 | 36.26 | 20.43 | 42.00 | 31.98 | **19.14** | 26.72 | 31.12 | 34.83 | **20.05** | 18.31 |
| AgeCutoff<65 | 62.05 | **67.81** | 68.57 | 73.94 | 76.16 | 55.07 | 65.28 | 72.34 | 75.45 | **59.01** | 57.63 |
| AgeCutoff>65 | 28.55 | 15.11 | 17.41 | 15.04 | **14.53** | 38.01 | 23.27 | 17.67 | **14.34** | 31.83 | 32.27 |
| AgeCutoff<80 | 84.62 | **79.83** | 82.64 | 86.24 | 88.08 | 84.94 | 83.90 | 86.42 | 86.91 | **84.14** | 83.14 |
| Zip or address | 100.00 | 100.00 | 100.00 | 100.00 | 100.00 | 100.00 | 100.00 | 100.00 | 100.00 | 100.00 | 100.00 |
| Medication | 83.51 | **74.31** | 79.63 | 81.52 | 84.88 | 86.27 | 81.00 | 81.66 | **81.00** | 84.11 | 83.73 |
| Diagnoses | 88.65 | **81.75** | 83.92 | 87.82 | 89.83 | 90.97 | 86.93 | 88.40 | **88.31** | 88.78 | 87.83 |
| Outpatient visits | 99.72 | 99.35 | **99.86** | 99.56 | 100.00 | 99.70 | 99.56 | 99.72 | **99.72** | 99.73 | 99.66 |
| Obs period 1 w | 1.83 | 2.63 | **0.35** | 2.16 | 0.58 | 2.67 | 2.41 | 0.85 | **0.69** | 2.09 | 2.17 |
| Obs period 2 w | 1.71 | 2.41 | **0.29** | 1.82 | 0.58 | 2.53 | 2.28 | 0.77 | **0.61** | 1.96 | 2.08 |
| Obs period 1 m | 1.53 | 1.97 | **0.24** | 1.48 | 0.58 | 2.32 | 1.97 | 0.62 | **0.47** | 1.78 | 1.86 |
| Obs period 6 m | 1.02 | 0.76 | 0.09 | 0.66 | **0.00** | 1.67 | 0.87 | 0.31 | **0.17** | 1.22 | 1.21 |
| Obs period 1 y | 0.91 | 0.49 | 0.06 | 0.48 | **0.00** | 1.51 | 0.59 | 0.24 | **0.12** | 1.09 | 1.03 |
| Obs period 2 y | 0.78 | 0.30 | 0.02 | 0.32 | **0.00** | 1.34 | 0.28 | 0.19 | **0.07** | 0.95 | 0.93 |
| Obs period 6 y | 0.70 | 0.17 | 0.01 | 0.24 | **0.00** | 1.21 | 0.19 | 0.17 | **0.06** | 0.85 | 0.81 |

Table A4. Available percentage of patients' data upon applying all the 19 filters to the All of Us dataset. The underlined and bolded text indicates the most affected known race by that filter, and the bold text indicates the most affected known ethnicity.

**Table A5**

| Filter | All | Asian | Black or African | Mixed | White | Other | Unknown |
| --- | --- | --- | --- | --- | --- | --- | --- |
| Alive | 93.03 | 95.23 | 95.86 | 95.03 | **92.89** | 95.44 | 91.20 |
| AgeSex | 100.00 | 99.99 | 100.00 | **99.88** | 99.98 | 99.99 | 100.00 |
| AgeCutoff>18 | 45.78 | 46.15 | 70.50 | 70.31 | **46.10** | 15.96 | 54.53 |
| AgeCutoff<21 | 16.51 | **7.57** | 16.81 | 23.92 | 17.00 | 2.94 | 15.17 |
| AgeCutoff<40 | 35.53 | **32.08** | 52.00 | 55.92 | 35.95 | 10.75 | 37.86 |
| AgeCutoff<65 | 44.95 | 45.73 | 69.82 | 69.90 | **45.25** | 15.86 | 53.39 |
| AgeCutoff>65 | 24.35 | **15.28** | 16.84 | 19.35 | 25.06 | 4.85 | 27.29 |
| AgeCutoff<80 | 45.78 | 46.15 | 70.50 | 70.31 | **46.10** | 15.96 | 54.53 |
| Zip or address | 29.51 | 35.49 | 57.46 | 40.03 | **28.80** | 41.64 | 32.11 |
| Medication | 73.37 | 72.44 | **65.48** | 69.09 | 73.61 | 67.78 | 70.12 |
| Diagnoses | 92.96 | 92.13 | **87.27** | 91.91 | 93.05 | 91.22 | 92.02 |
| Outpatient visits | 45.80 | 46.34 | **29.28** | 40.32 | 46.18 | 35.22 | 40.70 |
| Obs period 1 w | 44.47 | 45.17 | **28.41** | 38.76 | 44.84 | 34.09 | 39.10 |
| Obs period 2 w | 44.43 | 45.15 | **28.38** | 38.71 | 44.80 | 34.04 | 39.05 |
| Obs period 1 m | 44.37 | 45.09 | **28.38** | 38.71 | 44.74 | 34.04 | 38.94 |
| Obs period 6 m | 44.19 | 44.90 | **28.27** | 38.59 | 44.56 | 33.88 | 38.89 |
| Obs period 1 y | 44.06 | 44.70 | **27.96** | 38.53 | 44.42 | 33.74 | 38.74 |
| Obs period 2 y | 43.85 | 44.45 | **27.76** | 38.42 | 44.22 | 33.55 | 38.63 |
| Obs period 6 y | 43.25 | 43.49 | **26.60** | 37.61 | 43.63 | 32.75 | 38.01 |

Table A5. Available percentage of patients' data upon applying all the 19 filters to the UK Biobank dataset. The underlined and bolded text indicates the most affected known race by that filter.

**Figure A1**

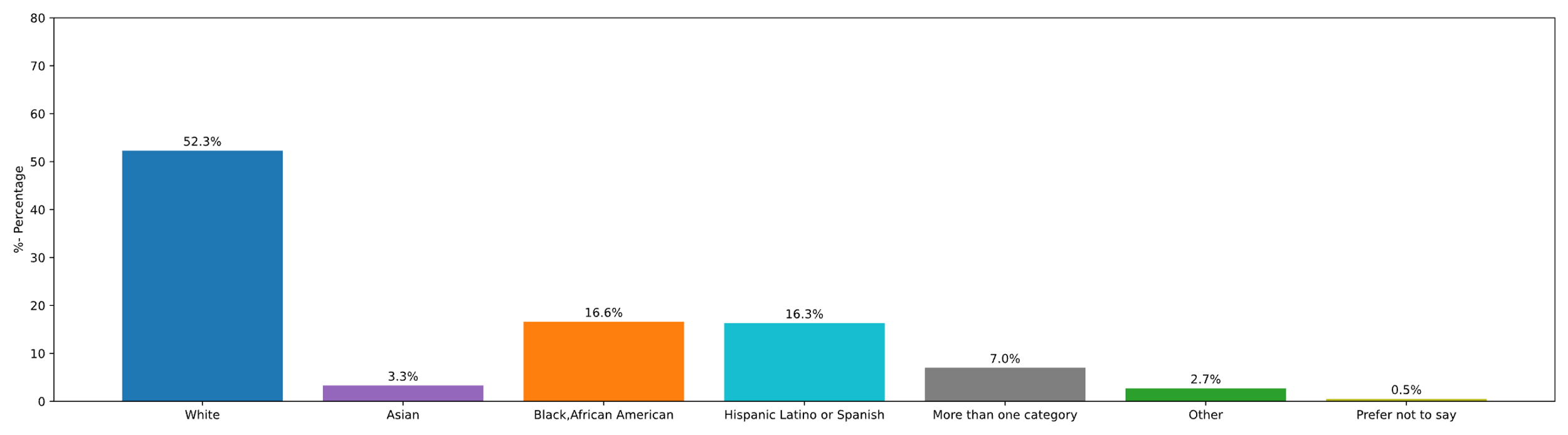

Figure A1. Distribution of the self-reported races and ethnicities of the participants who have completed the initial steps of the program, with or without EHR data(3).

**APPENDIX REFERENCES**

1.	www.census.gov [Internet]. United States Census Bureau. Available from: https://www.census.gov/quickfacts/fact/table/newyorkcitynewyork,losangelescountycalifornia,US/PST045223

2.	gov.uk [Internet]. Population of England and Wales. Available from: https://www.ethnicity-facts-figures.service.gov.uk/uk-population-by-ethnicity/national-and-regional-populations/population-of-england-and-wales/latest/

3.	All of Us Research Program. Researcher Workbench [Internet]. 2023. Available from: https://www.researchallofus.org/data-tools/workbench/
